# Diabetes and Stroke: Does an Excess Risk of Hemodynamic Cerebral Ischemia Exist?

**DOI:** 10.1101/2024.05.23.24307778

**Authors:** Christian Petitjean, Julien Labreuche

## Abstract

**BACKGROUND:** Diabetes is an independent risk factor for ischemic stroke. This study was undertaken to determine whether hemodynamic cerebral ischemia (HCI), which increases stroke severity, is more frequent in diabetic patients.

**METHODS:** Between 01/01/1990 and 30/06/2019, we revascularized 4117 carotid bifurcations in 3739 patients (2683 men and 1056 women; mean age: 71 (range: 28–97) years; 940 (25.1%) diabetics and 2799 (74.9%) nondiabetics). HCI was diagnosed clinically, under regional anesthesia, when a shunt needed to be inserted.

**RESULTS:** The HCI rate during carotid-clamping, requiring shunt placement, was 11.0% (114/1034) for diabetics and 7.1% (219/3083) for nondiabetics (odds ratio,1.62 [95% CI, 1.27–2.05]; *P*<0.0001).

**CONCLUSIONS:** Diabetes was associated with increased HCI risk in our patients with carotid stenosis. This HCI-associated excess risk might explain, in part, stroke severity in diabetics and heightened surgical risk. It might be prevented by HCI detection and modification of therapeutic management.

## INTRODUCTION

Embolic and hemodynamic mechanisms are the main causes underlying ischemic strokes of carotid origin.^1^ The hemodynamic cerebral ischemia (HCI) risk depends on the contribution of the contralateral internal carotid artery (ICA) and vertebral arteries via the circle of Willis, the ipsilateral external carotid artery via the ophthalmic artery and the leptomeningeal arteries.^2^ During carotid surgery, impaired collateral flow is associated with the need for shunt insertion.^3,4^

When HCI is present, cerebral perfusion is initially maintained by vasodilation of precapillary arterioles and the increased extraction coefficient of oxygen. Secondarily, vascular reserve exhaustion by degradation of arterial lesions engenders a loss of cerebral autoregulation, ischemic penumbra and cerebral infarction.^5^

Carotid revascularization with an incomplete circle of Willis enhances the postoperative ischemic stroke risk.^6^ The loss of cerebral autoregulation, attributable to HCI combined with ipsilateral carotid tight stenosis,^7^ heightens the risk of hyperperfusion and cerebral hemorrhage.^8–11^

Carotid occlusion is the primary cause of HCI.^4^ Carotid occlusions and tight stenoses lead to loss of cerebral autoregulation and cerebrovascular reserve,^11,12^ and have been associated with a 4-fold-increased stroke risk.^13^

Diabetes in an independent risk factor for ischemic stroke,^14,15^ whose associated mortality rate is higher^16^ and sequelae more serious^17,18^ than for nondiabetics. Diabetes increases the risk of stroke or death after surgical carotid revascularization or endoluminal angioplasty.^19^ It is, with contralateral ICA occlusion, 1 of the 7 factors doubling the stroke risk after carotid endarterectomy.^20^ Diabetes also enhances the cerebral hemorrhage risk associated with carotid surgery,^21^ thrombectomy^22^ or thrombolysis^23^ revascularization of the cerebral arteries.

This study was undertaken to examine whether the HCI frequency is higher in diabetics than nondiabetics and, if diabetes carries an excess HCI risk, whether it is independent of contralateral ICA occlusion.

## METHODS

### Study Population

Between 01/01/1990 and 30/06/2019, 4117 carotid bifurcations with atheromatous stenoses were revascularized in 3739 patients (2683 men and 1056 women; mean age: 71 (range: 28–97) years; 940 (25.1%) diabetics and 2799 (74.9%) nondiabetics) under regional anesthesia with a carotid-clamping test. The 49 operations without a clamping test were excluded: 27 operations under general anesthesia, 6 under regional anesthesia for evolving ischemic strokes requiring systematic shunting, 15 ICA occlusions revascularized and 1 major stroke before clamping test.

HCI was diagnosed clinically under regional anesthesia when consciousness perturbations, language difficulties and/or contralateral motor deficit appeared during the carotid-clamping test and required shunt placement.

### Statistical Analyses

Results are expressed as means±standard deviation for quantitative variables and number (percentage) for categorical variables. Bilateral ICA surgeries in a given patient (done in 378 patients) were considered independent events in all analyses. Patients’ main characteristics at the time of surgery are reported according to diabetes status and the magnitude of differences were assessed by calculating the standardized differences; an absolute standardized difference >20% was interpreted as meaningful.

The association between diabetes and HCI occurrence during carotid-clamping requiring shunt placement was assessed with a logistic-regression model before and after adjustment for meaningful differences between diabetic and nondiabetic patients’ characteristics; odds ratios (ORs) [95% confidence intervals (CIs)] were estimated from the logistic-regression model as effect size using nondiabetics as the reference. We further investigated that association according to presence or absence of contralateral ICA occlusion by entering the corresponding interaction terms into the logistic-regression models. Furthermore, univariable logistic-regression models were used to examine the association of diabetes severity according to diabetes subtype (insulin-dependent versus noninsulin-dependent) or glycemia level before surgery. Glycemia levels were analyzed as a continuous variable (after log-transformation to reduce skewness) and as a 4-level categorical variable (according to quartile distribution) to assess the form of the association. Statistical analyses were computed with a 2-tailed α-level of 0.05. Data were analyzed using SAS software version 9.4 (SAS Institute, Cary, NC).

## RESULTS

From January 1, 1990, to June 30, 2019, 3739 patients underwent 4117 ICA surgeries, under regional anesthesia, with a carotid-clamping test to treat atheromatous carotid-bifurcation stenosis. Among the 4117 ICA revascularizations, 1034 (25.1%) were done in diabetics and 3083 (72.9%) in nondiabetics. **Table 1** reports patients’ main characteristics at carotid-artery surgery, according to diabetes status. Their overall mean age was 71±10 years, 2978 (72.3%) were men and they did not differ according to diabetes status. As expected, body mass index was higher for diabetics, who were also more often hypertensive, hypertriglyceridemic and with coronary artery disease than nondiabetics (all meaningful standardized differences >20%). Among the 4117 ICA revascularizations, 1828 (44.4%) were done in patients with ipsilateral symptoms (580 cerebral transient ischemic attacks, 1061 minor strokes and 187 major strokes) and 200 (4.9%) in patients with contralateral ICA occlusion without difference according to diabetes status.

**Table 1.** Baseline Characteristics of Patients Undergoing Carotid Surgery According to Diabetes Status.

| Characteristic | Diabetes |  | Standardized |
| --- | --- | --- | --- |
|  | No (n=3083) | Yes (n=1034) | Difference, % |
| Age (years) | 71.2±10.6 | 70.9±9.2 | −2.5 |
| Men | 2197 (71.3) | 781 (75.5) | 9.7 |
| Body mass index (kg/m<sup>2</sup>)* | 25.2±3.7 | 27.3±4.2 | 53.3 |
| Location of lesions |  |  |  |
| Right side | 1549 (50.2) | 477 (46.1) | 8.2 |
| Left side | 1534 (49.8) | 557 (53.9) |  |
| History |  |  |  |
| Hypertension | 2166 (70.3) | 850 (82.2) | 28.4 |
| Hypercholesterolemia | 2034 (66) | 679 (65.7) | −0.7 |
| Hypertriglyceridemia | 934 (30.3) | 468 (45.3) | 31.2 |
| Smoker versus never smoker | 884/2940 (30.1) | 322/1001 (32.2) | 4.5 |
| Coronary artery disease | 837 (27.1) | 380 (36.8) | 20.7 |
| Neurological events |  |  |  |
| Neurological symptoms |  |  |  |
| Asymptomatic | 760 (24.7) | 272 (26.3) | 3.6 |
| Unspecific symptomatic sign | 366 (11.9) | 117 (11.3) |  |
| Specific symptomatic sign | 1957 (63.5) | 645 (62.4) |  |
| Ipsilateral symptoms |  |  |  |
| None | 1725 (56) | 564 (54.5) | 8.1 |
| Cerebral TIA | 447 (14.5) | 133 (12.9) |  |
| Minor stroke | 772 (25.0) | 289 (27.9) |  |
| Major stroke | 139 (4.5) | 48 (4.6) |  |
| Territorial infarct | 1060/2783 (38.1) | 376/939 (40.0) | 4.0 |
| Junctional infarct | 225/2903 (7.8) | 116/977 (11.9) | 13.9 |
| Contralateral ICA occlusion | 146 (4.7) | 54 (5.2) | 2.2 |

Values are mean±standard deviation or n/total N (%), unless stated otherwise.
\* 51 missing values (8 in the diabetes group).
Abbreviations: ICA, internal carotid artery; TIA, transient ischemic attack.

Overall, HCI appeared during 333 procedures (8.1%; [95% CI: 7.2–9.0%]). As shown in **Figure 1**, HCI frequency during carotid-clamping was significantly higher in diabetics (11.0%) than nondiabetics (7.1%), with an unadjusted OR of 1.62 [95% CI, 1.27–2.06]. After adjustment for baseline imbalance characteristics between the 2 groups, the difference remained unchanged (adjusted OR, 1.68 [95% CI, 1.30–2.16]).

**Figure 1.**
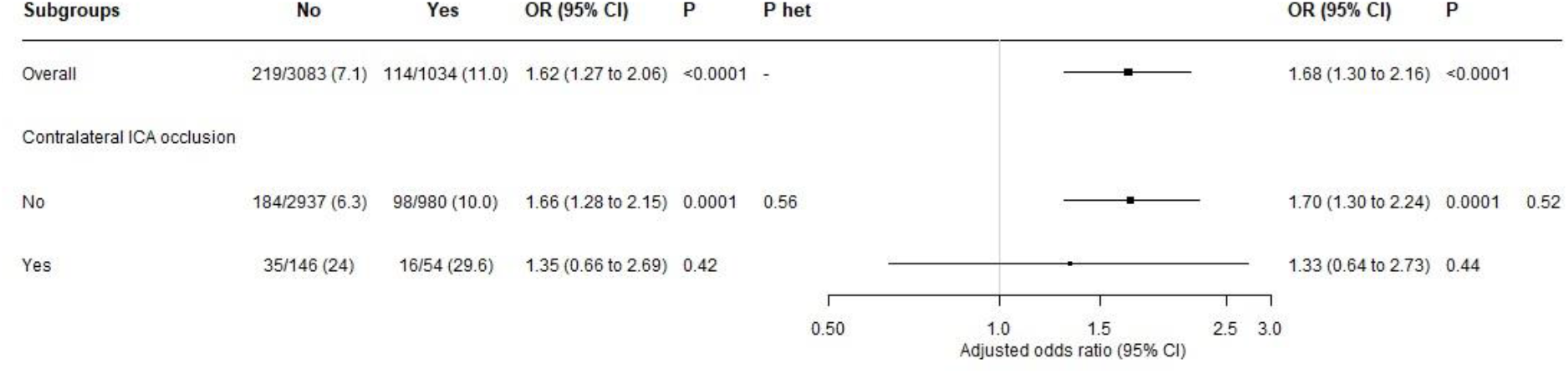
Association between diabetes and hemodynamic cerebral ischemia (HCI) rate, overall and according to contralateral internal carotid artery (ICA) occlusion status. Values are n/total N (%), unless stated otherwise. Odds ratio (ORs) were adjusted for body mass index, hypertension, hypertriglyceridemia and coronary artery disease. *P* het indicates *P*-value for heterogeneity in diabetes association and HCI rate according to contralateral ICA occlusion status. Abbreviations. CI, confidence interval.

Subgroup analyses according to the presence or absence of contralateral ICA occlusion showed that having a contralateral ICA occlusion had no impact on the association of diabetes and HCI rate (adjusted *P* for heterogeneity=0.52), with adjusted ORs of HCI for diabetics versus nondiabetics of 1.33 [95% CI, 0.64–2.73] with a contralateral ICA occlusion and 1.70 [95% CI, 1.30–2.24] without contralateral ICA occlusion (**Figure 1)**.

As shown in **Table 2**, diabetes severity assessed as insulin-dependent versus noninsulin dependent or the glycemia level before surgery were not associated with an increased HCI risk during carotid clamping.

**Table 2.**
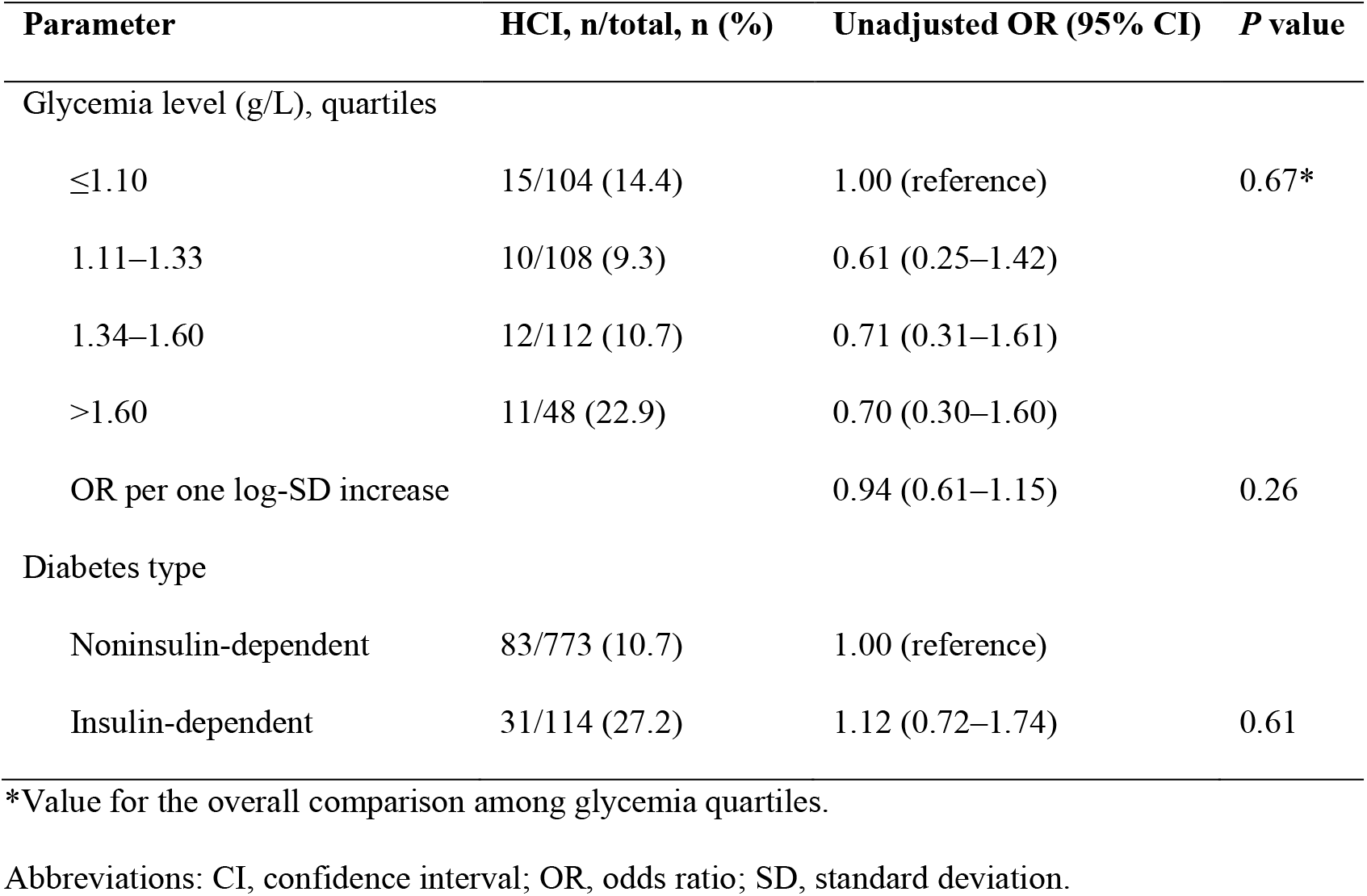
Association between Glycemia Level or Diabetes Type and the Hemodynamic Cerebral Ischemia (HCI) Rate among Diabetic Patients.

## DISCUSSION

The results of this study showed that diabetes was associated with enhanced HCI risk in patients with carotid stenosis. That risk was independent of contralateral ICA occlusion. It is likely linked to impaired collateral vascularization in diabetics^24,25^ and the concomitant decline of vascular reserve, but also to other factors increasing cerebral ischemia.^14,15,26,27^ In this study, when diabetes was associated with contralateral ICA occlusion, the HCI frequency was almost 4 times higher than when neither of those 2 factors was present (respectively: 35/146 (24%) and 184/2936 (6.3%)).

Intracranial arterial stenoses are more frequent in diabetics^25,26^ and associated with carotid stenosis, thereby increasing the stroke risk in patients treated medically but not those undergoing carotid surgery.^28^

### Diabetes and Antihypertensive Treatment

Hypertension is more frequent in diabetics,^29^ who are at greater risk of cardiovascular complications. Antihypertensive therapy significantly lowers that excess risk.^30–32^

Meta-analysis results led to the suggestion to base antihypertensive treatments on the cardiovascular risk rather than on blood-pressure measurement, while recommending selection of the individuals most likely to benefit.^33^ The finding of a subsequent analysis showed the benefit of antihypertensive therapy on diabetics’ stroke risk.^34^

Intensive blood-pressure control durably diminishes cerebral blood flow in diabetics with microvascular complications compared to those without and nondiabetics.^35^ Combining intensive antihypertensive therapy with intensive glycemia control lowered the stroke benefit in diabetics.^36^ Therefore, the diminished therapeutic benefit might be attributed to the impaired collateral vascularization, more frequent in diabetics, which would induce cerebral blood-flow hypoperfusion engendering less oxygen and sugar supplies, thereby favoring hypoxia and hypoglycemia.

### Diabetes and Carotid Surgery

Carotid surgery is beneficial in preventing ischemic strokes, regardless of whether tight carotid stenosis is symptomatic or asymptomatic.^37–39^ But that benefit is reduced by the increased stroke risk in diabetics.^40^ According to the randomized North American Symptomatic Carotid Endarterectomy Trial (NASCET), diabetes significantly doubled the perioperative stroke risk or death, but strokes were not analyzed independently of deaths.^20^ The results of the randomized Asymptomatic Carotid Atherosclerosis Study (ACAS) showed that the postoperative ischemic stroke risk was more than doubled in diabetics than nondiabetics (respectively, (3.9% versus 1.4%; *P*<0.05).^41^ The excess stroke risk of carotid surgery in diabetics disappeared when HCI was detected under regional anesthesia and prevented by shunt insertion.^12,42^ HCI caused a loss of vascular reserve that increases the ischemic stroke risk^13,43^ and loss of cerebral autoregulation that enhances the risk of cerebral hemorrhage and convulsions.^8–11^

In addition, diabetes increases mortality after cerebral hemorrhage.^44^ That excess risk has only rarely been demonstrated in diabetics,^21^ perhaps because of the rarity of this complication. In our series, surgery on diabetics with a reperfusion risk represented only 1.5% (62/4117) of all operations.

### Stroke Prevention in Diabetics

Analysis of vascular supply, cerebral autoregulation and vascular reserve is decisive in managing strokes, whether the patient is diabetic or not.^45^ That analysis could contribute to managing hypertension by enabling detection of hypertensive patients with altered vascular reserve, so as to moderate antihypertensive and glycemia-control therapies in this subgroup and by maintaining intensive antihypertensive treatment and intensive hypoglycemia treatments in patients with normal vascular reserve.

These surgical patients at risk can be identified during the preoperative work-up,^2,3 46,47^ during the intervention under general anesthesia by transcranial evaluation of oxygen saturation^48^ or under regional anesthesia with a clamping test.^8^ That detection prevents intraoperative hemodynamic ischemic strokes by the selective insertion of a shunt^8,12^ and cerebral hemorrhages by controlling blood pressure after surgery^49^ or endoluminal angioplasty.^50^

When carotid stenosis is asymptomatic, especially when the contralateral ICA is occluded, the loss of vascular reserve could be an additional criterion supporting surgery—if it is a tight stenosis—or close monitoring for stenoses just below the limit indicating surgery, whether the patient is diabetic or not.

### Limitations

This study concerned only diabetics with carotid stenosis. The latter is associated with a heightened frequency of intracranial artery stenoses^51^ that favors HCI.^24^ Our results must be verified for diabetics without carotid stenosis. Moreover, HCI is only one of the factors worsening strokes in diabetics and its role has not been clearly evaluated. In addition, glycemia was not systematically assessed immediately preoperatively.

## CONCLUSIONS

Diabetes increased the HCI frequency in patients with carotid stenosis, which could explain, in part, the severity of strokes in diabetics and the excess risk of carotid revascularization. Taking that excess risk into account might contribute to lowering stroke frequency and severity. HCI should be considered one of the cardiovascular risk factors used to manage hypertension in diabetics.

## Data Availability

All data produced in the present study are available upon reasonable request to the authors

## Non-standard Abbreviations and Acronyms

CI: confidence interval
HCI: hemodynamic cerebral ischemia
ICA: internal carotid artery
OR: odds ratio

## Acknowledgments

We thank Mylène Redondi for database management and Janet Jacobson for editorial assistance.

## Conflicts of Interest

The authors declare no conflict of interest.

## Notes

### Competing Interest Statement

The authors have declared no competing interest.

### Funding Statement

This study did not receive any funding

### Author Declarations

Ethics committee/IRB of Institut Mutualiste Montsouris gave ethical approval for this work

